# An Alternative Approach to Tablet Splitting and Grinding for Medication Administration

**DOI:** 10.1101/2022.08.26.22279282

**Authors:** Rajneesh Taneja, Joseph Scarim, Poonam G Pande, Anthony Scarim, Milap C Nahata, Rita K Jew, Koteswara Inabathina

**Author notes:** Correspondence Poonam Pande, TB Alliance, 40 Wall Street, 24^th^ Floor, New York, NY 10005.

## Abstract

**Purpose:** Tablet formulations fail to meet the needs of patients unable to swallow tablets such as pediatric, elderly, and patients that must receive medications via feeding tubes. Our aim was to develop and test a new, simple device (XTEMP-R™) and the methodology for converting tablets into a homogeneous suspension for medication administration.

**Methods:** We developed a new device comprised of a flexible receptacle, a tight-fitting cap, and a suction cup bottom to convert tablets into liquid preparations. TB treatment drugs, TBAJ-876 and TBI-223, were dispersed within the device utilizing water and commonly available suspending vehicles. We investigated the effectiveness of the XTEMP-R device in dispersing tablets. This was accomplished by visual observations, determining the fineness of dispersion, and measuring the total drug recovery from the dispersions in XTEMP-R. We investigated the accuracy and reproducibility of delivering aliquots from these suspensions by determining the dose reproducibility upon suspension and upon redispersion after 24 hours. The effectiveness of the device was also evaluated using commercially available tablets of acetaminophen, amlodipine, glimepiride, metformin and valsartan.

**Results:** The suspensions were visually uniform without any large particles. The suspensions passed through #18 sieve confirming that the particles were less than 1000 μm. Average total dose recovery of three suspensions each was determined to be 101.3% and 99.2% for TBI-223 and TBAJ-876, respectively. Reproducibility from aliquots of 2 mL each were 98.9-99.7% for three replicates of TBI-223 suspensions, and 102.6-103.2% for TBAJ-876 suspensions. Aliquots tested after 24 hours confirmed uniform re-dispersibility.

**Conclusion:** We have demonstrated that XTEMP-R can be utilized to prepare homogeneous suspensions can be prepared conveniently and efficiently in less than 10 minutes without any drug loss. Aliquots for partial dose delivery can be withdrawn accurately. These findings demonstrate that XTEMP-R can be used to accurately deliver doses of suspensions for patients who cannot swallow tablets.

## Introduction

Tablets remain the most common modality for oral dose administration of drugs. They represent 28 of the top 30 prescription drugs in 2021 [1]. However, these formulations are not suitable for some patients, especially young children, and those suffering from dysphagia. Tablet administration is also a challenge for patients receiving enteral tube feeding [2]. If an alternate oral formulation or route of administration is unavailable, manipulation of the dosage form may be undertaken at the point of administration to provide the prescribed dose [3]. Manipulation involves physical alteration of the dosage form, including tablet splitting, crushing, or grinding [4]. Dose adjustment is commonly achieved through tablet splitting, whereas tablet crushing or grinding aid the swallowing or administration via an enteral feeding tube [4].

Tablets are often pulverized (crushed or ground) with a mortar and pestle or other tablet grinding device to facilitate administration [2,5]. However, this process is tedious and often leads to the loss of the prescribed dose during preparation or administration [5]. In one study, it was demonstrated that two rinses of the grinding devices with water were required to prevent significant drug loss [5]. Screwcap crushers yield larger particles that are not suited for feeding tube administration [4]. Also, some hazardous drugs may pose risks to caregivers preparing the doses through the inhalation of generated powder containing the drug, or through contact with skin, nose, and eyes [4].

Another common practice is tablet splitting [6,7]. It is used for many reasons including delivering partial dose of the tablet, facilitating swallowing, or cost savings [8,9]. However, this process raises the issues of drug loss, variation in dose uniformity, dose accuracy, and weight uniformity between fragments [8]. Many tablets may not be suitable for splitting due to factors such as hardness, size, shape, or absence of score line.

Steadman and colleagues [5] called for a device that minimizes drug loss and maximizes medication delivery, i.e., a device that ensures a sealed environment for crushing followed by mixing with a fluid and subsequent consumption from the same receptacle [5]. Salmon and colleagues [2] expressed a need for a combined crushing and suspending device.

TBAJ-876 (US IND number 145,349) and TBI-223 (US IND Number 134,334) are two new chemical entities under development as immediate-release tablets for the treatment of tuberculosis. The tablets are available in 100 mg and 600 mg strengths, respectively. Dose titration is required while conducting early clinical studies with these drugs. A liquid formulation allows for the flexibility of aliquoting the desired dose in any fraction. The alternative is to manufacture tablets in multiple strengths and choose a combination of different strength tablets for the desired dose.

The objective of our work was to develop a device and methodology that enabled the dispersion of TBI-223 and TBAJ-876 immediate-release tablets into a suspension in water without the need for crushing or grinding in a contained environment. Further, a suspending vehicle can be added to make it into a homogenous suspension that allows for accurate partial dose delivery.

A new, simple device (XTEMP-R™) was developed, and the methodology for converting tablets into a homogeneous suspension with this device is described in this paper. This device and methodology can be used in any setting without crushing, grinding, or splitting the tablet dosage form. Flavors and sweeteners may be added to the suspension for enhancing palatability. Our work is not suited for modified release, delayed-release, or controlled-release tablets and capsules. In this paper, the term tablet refers to immediate-release tablet. The term dispersion refers to the disintegration of the tablet in a liquid media resulting in a suspension. The terms dispersion and suspension convey the same meaning.

To demonstrate the utility of the XTEMP-R device, we investigated (a) time to disperse the tablets, (b) fineness of dispersion, (c) total dose recovery, and (d) accuracy of delivering aliquots from suspensions.

In addition to TBAJ-876 and TBI-223, the dispersibility of a few commercial tablets was also conducted to verify the utility of this device.

## Materials and methods

### Drugs and vehicles

TBI-223 IR tablets, 600 mg, TB Alliance, lot 6767AU01, retest 11/2022

TBAJ-876 Tablets, 100 mg, TB Alliance, Batch 6767BG01, retest 10/2022

Valsartan tablets, USP, 320 mg (Macleods, lot EVI2032A, exp 07/2022)

Tylenol® (acetaminophen) extra-strength tablets, 500 mg (Johnson & Johnson, lot AFA111, exp 05/2025)

Metformin tablets, USP, 1000 mg (Ascend, lot 4200094A, exp 07/2023)

Glimepiride tablets, USP, 2 mg (Accord, lot P2005847, exp 10/2023)

Amlodipine tablets, USP, 2.5 mg, (Ascend, lot 21140696, exp 01/2024)

ORA-Blend®, Perrigo, lot 2077398, exp. 28 Feb 2025

ORA-Sweet®, Perrigo, lot 1346049, exp 31 July 2024

Water, Distilled, Culligan, R22075 06:56 L1 Best By 03/2023

### Chemicals

Acetonitrile, HPLC, Spectrum Chemical Mfg. Corp, item HP412, lot 20020323

Water, HPLC, Concord Technology, 8003LC4000, lot 201222

Ammonium Acetate, Sigma-Aldrich, item 73594-25G-F, lot BCCH0197

Trifluoroacetic acid (TFA), HPLC grade, Sigma-Aldrich, item 91707-10X1ML, lot bccg6537

Sodium lauryl sulfate (SLS), Sigma-Aldrich, Reagent grade, item 436143, lot MKCQ6608

### Equipment and supplies

HPLC: Hitachi L-2100 pump, Shimadzu SPD-10AVP detector, Shimadzu SCL-10AVP controller, Shimadzu CTO-AVP column heater, SRI Instruments PeakSimple chromatography data system Column: Zorbax Bonus RP (C18, 4.6 mm x 150 mm, 3.5μm)

Column: Waters Symmetry Shield RP-8 (4.6 mm x 150 mm, 5μm)

Syringe Filters, Tisch Scientific, Glass Fiber, Hydrophilic, 25mm, 1.0um, PN SF14699, lot 170420083

Syringe Filters, PALL Life Sciences, PVDF, 13mm, 0.45um, PN 4545, lot A10648946

### XTEMP-R™ device

XTEMP-R comprises a flexible receptacle, a tight-fitting cap, and a suction cup bottom (Fig 1). It is a soft walled device manufactured with biocompatible, low hardness HCR (high-consistency silicone rubber) sourced from Wacker, Germany. The construction material, platinum-cured silicone, complies with FDA 21 CFR 177.2600, German Bfr XV, France Arrete Du 25, ROHS, TSE/BSE free, USP Class VI Certification, and ISO 10993 Certification. These devices are produced by a compression molding process (Ami Polymers Pvt. Ltd., India) in a class 10000 cleanroom. The height of the XTEMP-R device is 140 mm, the internal diameter is 22 mm, and the capacity is approximately 40 mL. The device has volume markings on the outside at 5-mL intervals starting at 15 mL. This device is resistant to extremes of environments and temperatures and is designed to be leak-resistant and is easy to squeeze between the fingers for manual dispersion of the tablets.

**Fig 1.**
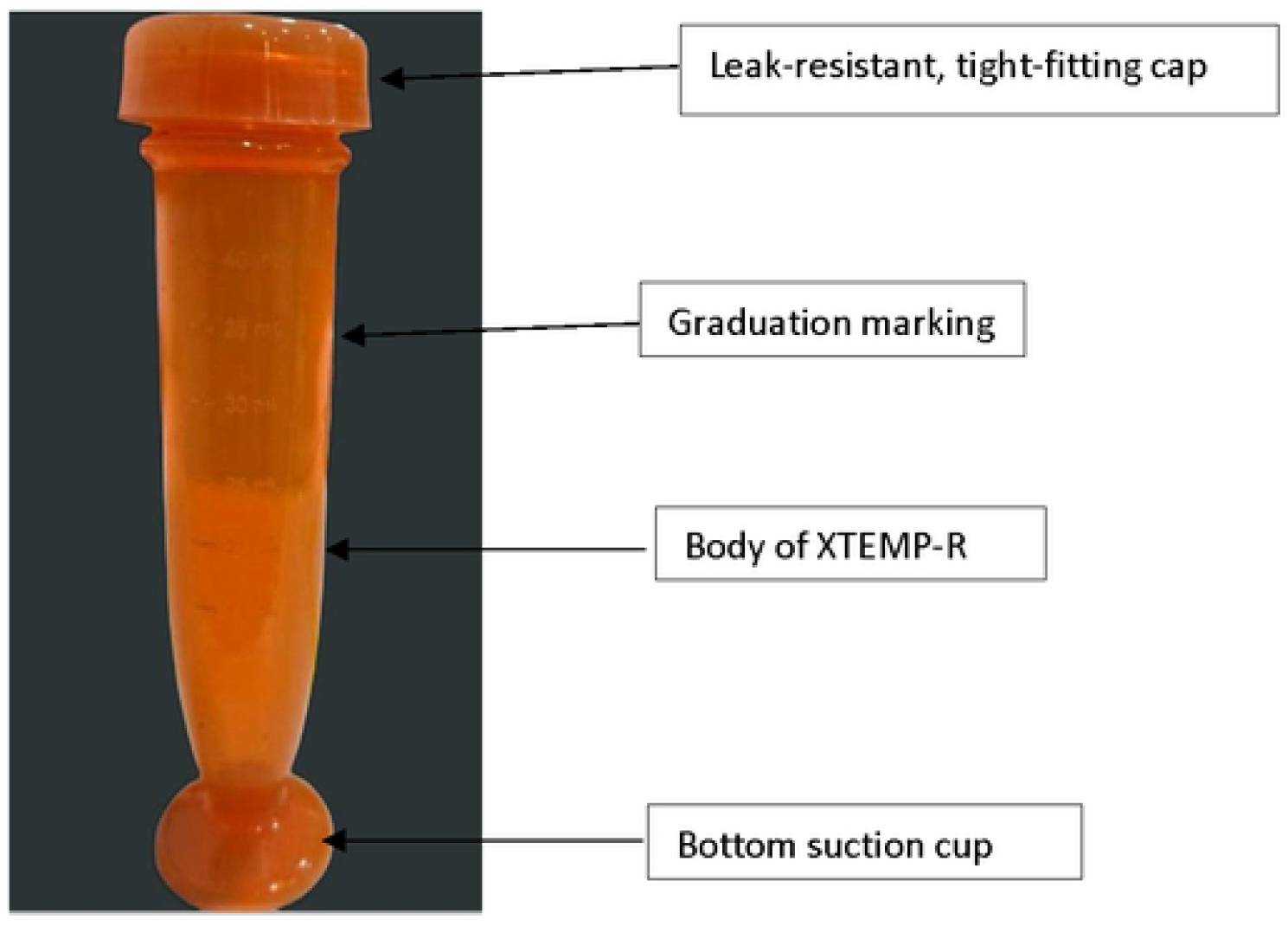
X-TEMP-R™ Device.

### Preparation of suspensions from tablets using XTEMP-R

The general preparation technique is displayed in Fig 2 with preparation of TBI-223 suspension as an example.

**Fig 2.**
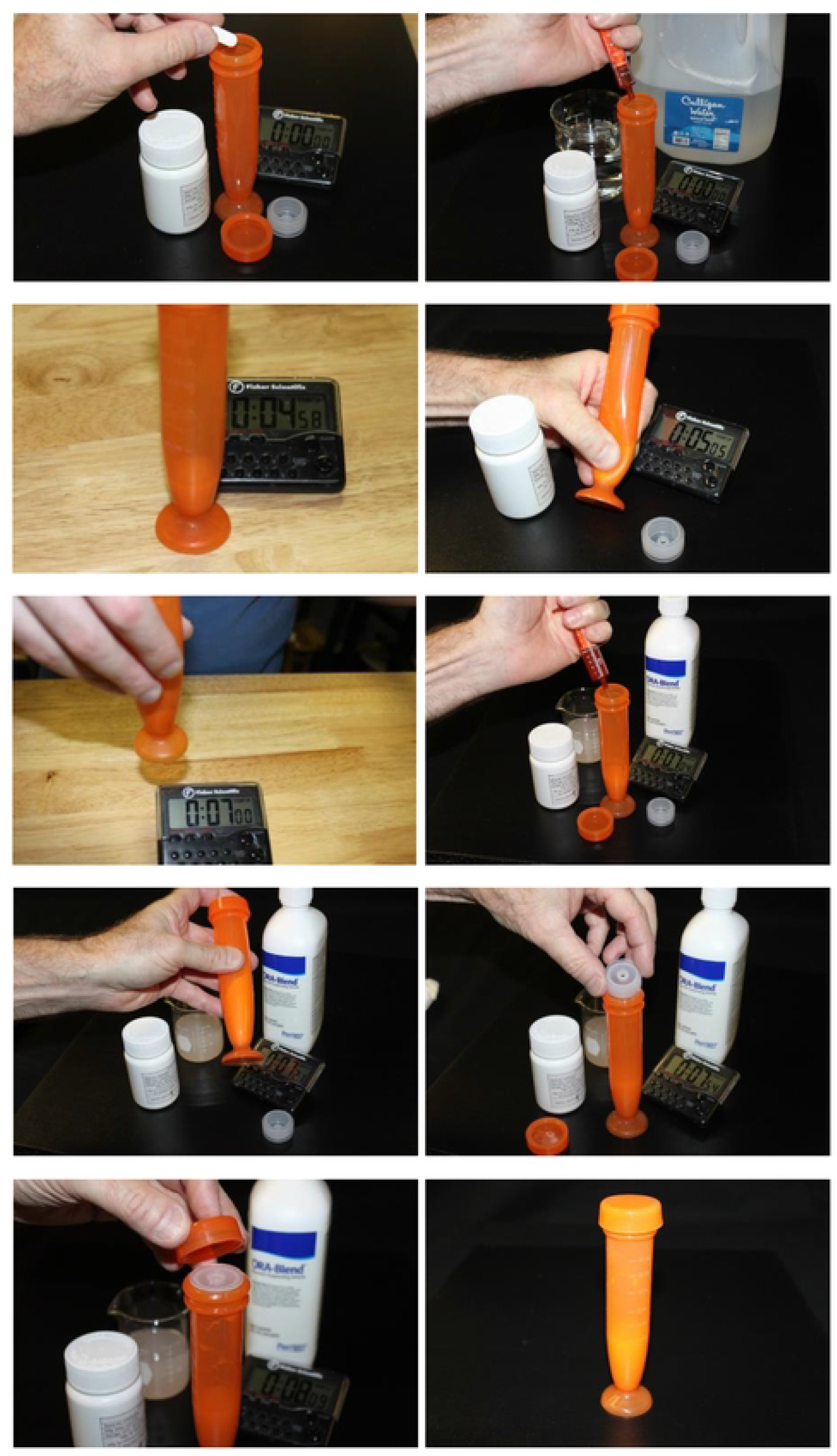
Preparation of TBI-223 Suspension from Tablets.

#### TBAJ-876

One tablet of TBAJ-876 100 mg was added to the XTEMP-R followed by 5 mL of water. The device was capped and swirled for 5 seconds and left undisturbed for 5 minutes. Then the tablet was manually massaged in the XTEMP-R for 2 minutes. ORA-Sweet suspending vehicle, 19.5 mL was added to the dispersion in the XTEMP-R make a final volume that coincided with the 25-mL mark on the device and a final concentration of 4 mg/mL. The device was capped and shaken 25 times to mix thoroughly. For redispersion, the suspension was shaken 25 times.

#### TBI-223

One tablet of TBI-223 600 mg was added to the XTEMP-R followed by 5 mL of water. The device was capped and swirled for 5 seconds and left undisturbed for 5 minutes. Then the tablet was manually massaged in the XTEMP-R for 2 minutes. ORA-Blend suspending vehicle, 9.4 mL was added to the dispersion in the XTEMP-R to make a final volume that coincided with the 15-mL mark on the device and a final concentration of 40 mg/mL. The device was capped and shaken 25 times to mix thoroughly. For redispersion, the suspension was shaken 25 times.

### Evaluation of suspensions prepared in XTEMP-R

#### Visual observation

##### TBAJ-876 and TBI-223

The prepared suspensions of TBAJ-876 and of TBI-223 were shaken, and then an aliquot of each was poured onto a watch glass. The suspensions were examined for color, uniformity, and presence of large particles, agglomerates, or clumps.

#### Fineness of dispersion

Dispersions of tablets in water prepared in the XTEMP-R device were tested for fineness of dispersion.

##### TBAJ-876 and TBI-223

One tablet was added to the XTEMP-R followed by 5 mL of water. The tablet was allowed to soak for 5 minutes, followed by 2 minutes of massaging in the XTEMP-R. The dispersion was poured through a #18 sieve (1000 μm opening) with a pan at the bottom for collecting the dispersion. This test was performed in triplicate by each of two analysts.

#### Total dose recovery from tablet dispersion in XTEMP-R

##### TBAJ-876

The prepared 25 mL suspension in the XTEMP-R device was shaken 25 times and transferred to a 250 mL volumetric flask. The device was rinsed with 15 mL of water and the rinse was added to the volumetric flask with the suspension. This stock solution was diluted to volume with 1:1 acetonitrile:water. A sample of this stock solution was further diluted with diluent (Table 1) to a concentration of 0.04 mg/mL. An aliquot of this diluted sample was filtered and analyzed by HPLC as described under *Analytical test methods* below. The recovery was tested on three preparations.

**Table 1.** Details of Stability-Indicating HPLC Methods for Potency Testing of TBI-223 and TBAJ-876 Suspensions.

| Parameter | Details |  |
| --- | --- | --- |
|  | TBI-223 | TBAJ-876 |
| Instrument | Hitachi L-2100 pump, Shimadzu SPD-10AVP detector, Shimadzu SCL-10AVP controller, Shimadzu CTO-AVP column heater, SRI Instruments PeakSimple chromatography data system |  |
| Column | Zorbax Bonus RP (C18, 4.6 mm x 150 mm, 3.5µm) | Waters Symmetry Shield RP-8 (4.6 mm x 150 mm, 5µm) |
| Wavelength | 258 nm | 236 nm |
| Column Temp. | 30 °C | 40 °C |
| Flow Rate | 0.8 mL/min | 2.0 mL/min |
| Injection Volume | 10 µL | 5 µL |
| Syringe filter | 1.0 µm glass fiber filter | 0.45 µm PVDF syringe filter |
| Standard and sample Concentration | 0.05 mg/mL | 0.04 mg/mL |
| Mobile Phase | Mobile Phase-A:<br>0.05% TFA in water<br>Mobile Phase-B:<br>0.05% TFA in Acetonitrile | 0.01M Ammonium acetate:<br>Acetonitrile (1:3) |
| Diluent | 0.2% SLS in Water:Acetonitrile (1:1) | Water:Acetonitrile (1:4) |
| Run Time | 8 minutes<br>(Gradient: 0.0-2.0 mins: A:64% and B:36%; 2.0-4.5 mins: A:64% and B:36% to A:46% and B:54%; 4.6-8.0 mins: A:64% and B:36%) | 7 minutes<br>(Isocratic) |
| Peak retention time | About 3.6 minutes | About 4.4 minutes |
| Integration | Peak area method using PeakSimple data acquisition system | Peak area method using PeakSimple data acquisition system |

##### TBI-223

The prepared 15 mL suspension in the XTEMP-R device was shaken 25 times and transferred to a 250 mL volumetric flask. The device was rinsed with 15 mL of water and the rinse was added to the volumetric flask with the suspension. A volume of 30 mL of acetonitrile was added to the flask and mixed. This stock solution was diluted with diluent (Table 1) to volume. A sample of this stock was further diluted with diluent to a concentration of 0.05 mg/mL. An aliquot of this sample was filtered and analyzed by HPLC as described under *Analytical test methods* below.

The recovery was tested on three preparations.

#### Dose reproducibility of aliquots of tablet dispersion in XTEMP-R

A general technique for withdrawal of aliquots for partial doses is represented in Fig 3.

**Fig 3.**
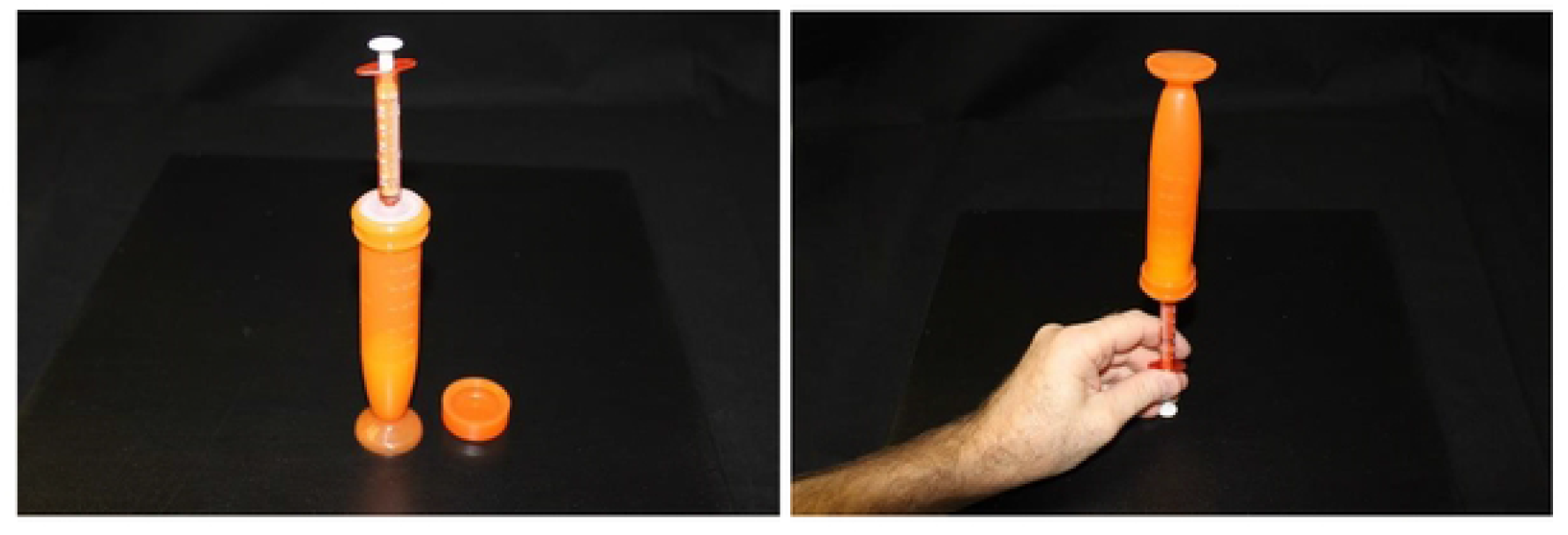
Withdrawal of an Aliquot from XTEMP-R for Partial Dose of TBI-223 Suspension.

##### TBAJ-876

Prior to withdrawal of the dose aliquot, a syringe adapter was attached to the opening of XTEMP-R and the device was capped. The prepared 25 mL suspension in the XTEMP-R device was shaken 25 times and a 3-mL oral syringe was attached to the syringe adapter. A 2-mL aliquot was removed for an 8-mg dose. The dose reproducibility was tested on three 2-mL aliquots withdrawn from the same preparation. For each withdrawal of the aliquot, the capped device was shaken to simulate the procedure that would be followed by the patient or caregiver. This was repeated on three different preparations. Each 8-mg dose was diluted with diluent to a concentration of 0.04 mg/mL, filtered, and analyzed by HPLC. The remaining suspension in the XTEMP-R was left undisturbed on the bench for about 24 hours. The preparations were redispersed by shaking and one aliquot from each preparation was withdrawn and tested.

##### TBI-223

Prior to withdrawal of the dose aliquot, a syringe adapter was fitted on to the device and the device was capped. The prepared 15 mL suspension in the XTEMP-R device was shaken 25 times and a 3-mL oral syringe was attached to the syringe adapter. A 2-mL aliquot was removed for an 80-mg dose. The dose reproducibility was tested on three 2-mL aliquots withdrawn from the same preparation. For each withdrawal of the aliquot, the capped device was shaken to simulate the procedure that would be followed by the patient or caregiver. This was repeated on three different preparations. Each dose withdrawn was transferred into a 100-mL volumetric flask. Acetonitrile 2 mL was added to the flask and swirled to mix the sample with the solvent.

The mixture in the flask was then diluted with diluent to volume. An aliquot of 3 mL of this stock sample preparation was diluted to 50 mL to a concentration of 0.05 mg/mL. An aliquot of this sample preparation was filtered and analyzed by HPLC. The remaining suspensions in the XTEMP-R were left undisturbed on the bench for about 24 hours and an aliquot from each preparation was tested after redispersing.

#### Analytical test methods for TBAJ-876 and TBI-223

Stability-indicating validated HPLC methods were used for the potency testing of TBI-223 and TBAJ-876 suspensions. Details of the HPLC methods are provided in Table 1.

##### TBAJ-876 quantification method validation

The method was validated for specificity (placebo interference and forced degradation), linearity, filter study, precision (system suitability and repeatability), and accuracy according to International Conference on Harmonization (ICH) guidelines [10]. Specificity was assessed to verify the absence of interference from tablet excipients, and suspension matrix and to establish the analyte peak does not interfere with the possible degradants of the forced degradation of samples. Seven-point calibration curve was constructed with standard solutions in the range of 50% (20 μg/mL) to 150% (60 μg/mL) of standard concentration i.e., 40 μg/mL.

##### TBI-223 quantification method validation

The method was validated for specificity (placebo interference and forced degradation), linearity, filter study, precision (system suitability and repeatability), and accuracy according to ICH guidelines. Specificity was assessed to verify the absence of interference from tablet excipients, and suspension matrix and to establish the analyte peak does not interfere with the possible degradants of the forced degradation of samples. Five-point calibration curve was constructed with standard solutions in the range of 50% (25 μg/mL) to 150% (75 μg/mL) of standard concentration i.e., 50 μg/mL.

### Dispersions of commercial tablets prepared in XTEMP-R

We tested five different commercially available tablets to determine effectiveness of the XTEMP-R device in producing a fine dispersion that passed through a #18 sieve (1000 um opening). The commercial tablets were selected to represent small and large tablets.

Manufacturer details, composition, tablet weight and drug content of the commercial tablets tested are provided in Table 2. Metformin tablets were included for evaluation to challenge the dispersing functionality of the XTEMP-R device, as they represent high dose, and high drug load tablets with a high drug to excipient ratio.

**Table 2.** Commercial Tablets Tested for Dispersion Times for Fineness of Dispersion.

| Product | Manufacturer | NDC | Inactive Ingredients | Tablet Weight (mg) | % Drug in Tablet |
| --- | --- | --- | --- | --- | --- |
| Valsartan 320 mg | MacLeods | 33342-065-10 | Colloidal silicon dioxide, crospovidone*, hypromellose, iron oxides (yellow, black and/or red), microcrystalline cellulose, magnesium stearate, polyethylene glycol, talc and titanium dioxide | 1030 | 31.1 |
| Acetaminophen 500 mg (Tylenol®) | McNeil | 50580-449-96 | Carnauba wax, corn starch, FD&C red no. 40 aluminum lake, hypromellose, magnesium stearate, modified starch, polyethylene glycol, powdered cellulose, pregelatinized starch, propylene glycol, shellac, sodium starch glycolate*, titanium dioxide | 610 | 82.0 |
| Glimepiride 2 mg | Accord | 16729-002-01 | Lactose monohydrate, sodium starch glycolate*, povidone, and magnesium stearate. Ferric oxide yellow and FD &C blue #2 aluminum lake | 170 | 1.2 |
| Amlodipine 2.5 mg | Ascend | 67877-197-90 | Microcrystalline cellulose, dibasic calcium phosphate anhydrous, sodium starch glycolate*, colloidal silicon dioxide and magnesium stearate | 100 | 2.5 |
| Metformin 1000 mg | Ascend | 67877-563-01 | Povidone (K-30), povidone (K-90), pregelatinized starch*, and magnesium stearate. Coating: artificial blackberry flavor, hypromellose and polyethylene glycol. | 1080 | 92.6 |

\*disintegrant

We tested the dispersibility of these commercial tablets by using the same methodology of soaking them in water for 5 minutes and massaging for 2 minutes, similar to the TBAJ-876 and TBI-223 tablets. The suspensions were tested for fineness of dispersion.

We also tested these commercial tablets for the shortest massaging time that could accomplish dispersion for fineness. One tablet was added to the XTEMP-R followed by 5 mL of water and massaged in the XTEMP-R device until the analyst felt that the tablet was completely dispersed, and the time was recorded. The suspension was poured through a #18 sieve (1000 μm opening) with a pan at the bottom for collecting the dispersion. This test was performed in triplicate by each of two analysts.

## Results

### Preparation of suspensions from tablets using XTEMP-R

#### TBAJ-876 and TBI-223

The suspension preparation technique for the two tablets, TBAJ-876 and TBI-223, was found to be simple, efficient and could be performed well in this closed system. The suspensions were prepared in less than 10 minutes without any complicated, tedious, or messy steps. The details of the suspension evaluation are provided below.

### Evaluation of suspensions prepared in XTEMP-R

#### Visual observation

##### TBAJ-876 and TBI-223

Fig 4 shows the visual appearance of the suspensions of the two drugs, TBI-223 and TBAJ-876. Both suspensions had uniform appearance and the particles were observed to be homogeneously dispersed. TBI-223 suspension was observed to be milky white, opaque, and uniform when light was shown from below the watch glass containing the suspension. TBAJ-876 suspension was translucent with distinct particles visible and uniformly dispersed. The appearance of the suspension as translucent or opaque is dependent on the ingredients and composition of the tablets and the dispersion vehicle.

**Fig 4.**
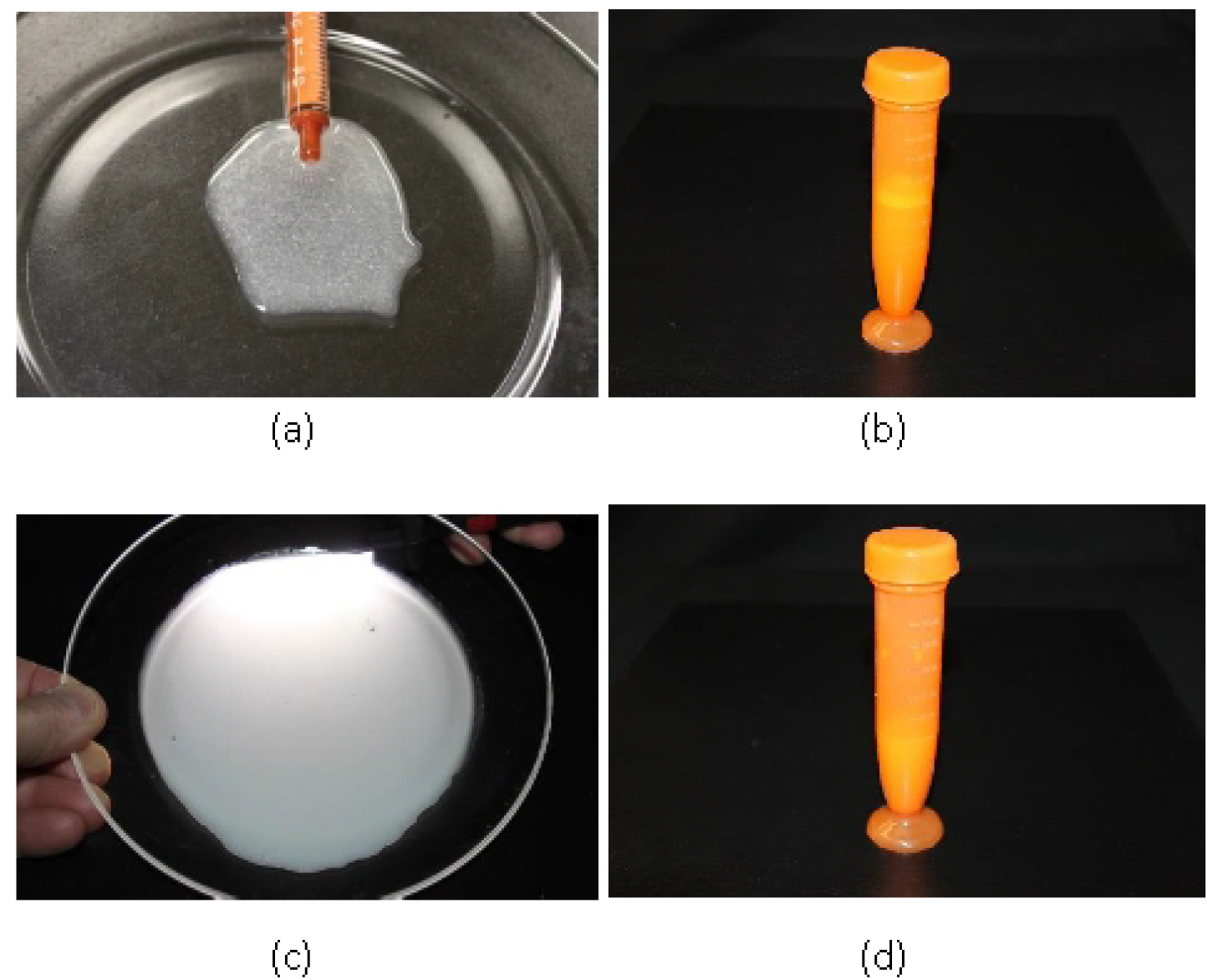
Visual Observation of Suspensions Prepared in XTEMP-R. (a) Appearance of TBAJ-876 suspension, (b) TBAJ-876 suspension in XTEMP-R device, (c) Appearance of TBI-223 suspension, (d) TBI-223 suspension in XTEMP-R device.

### Fineness of dispersion

#### TBAJ-876 and TBI-223

The dispersions prepared with 5-minute soaking and 2-minute massaging times in the XTEMP-R device passed through the #18 sieve. No particles were retained indicating that all particles were below 1000 μm and dispersed particles were not forming aggregates or agglomerates.

### Total dose recovery from tablet dispersion in XTEMP-R

#### TBAJ-876 and TBI-223

The total dose recovered from the triplicate preparations of TBAJ-876 and TBI-223 in XTEMP-R are presented in Table 3. The recovery of TBAJ-876 from three distinct preparations in XTEMP-R ranged from 98.8-100.0% with an average recovery of 99.2%. The recovery of TBI-223 from three distinct preparations in XTEMP-R ranged from 100.4-102.9% with an average recovery of 101.3%. These ranges include inherent tablet-to-tablet content variability as well as analytical variability.

**Table 3.**
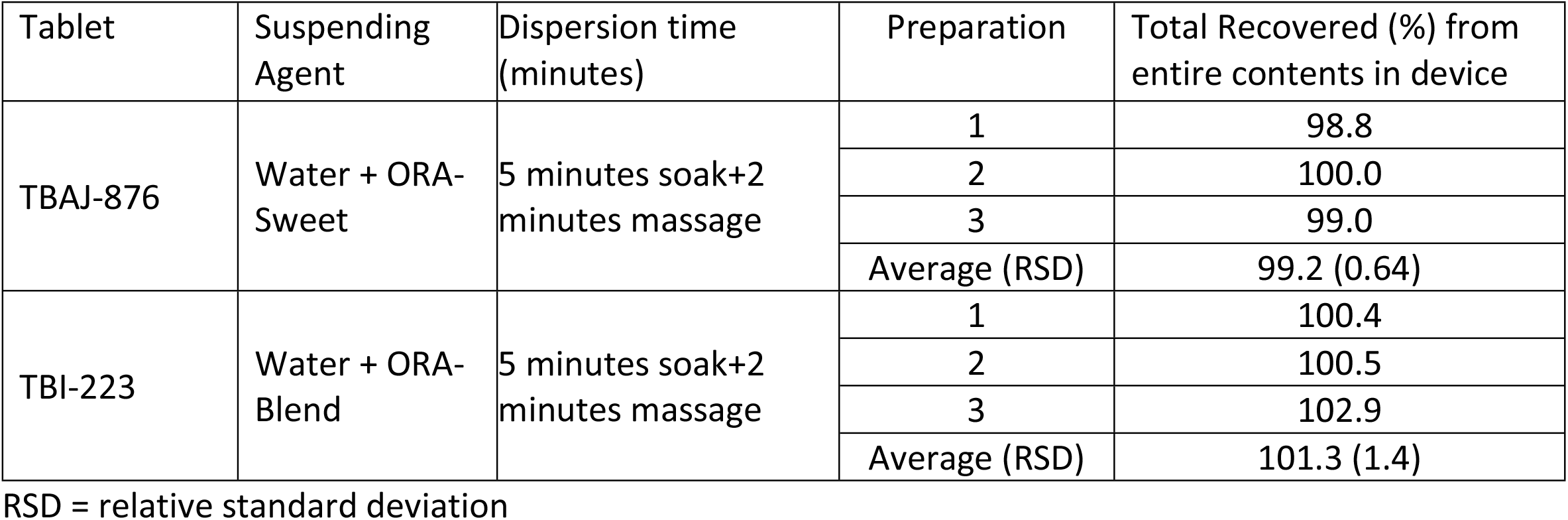
Recovery of TBI-223 and TBAJ-876 from Suspensions Prepared in XTEMP-R.

### Dose reproducibility of aliquots of tablet dispersion in XTEMP-R

#### TBAJ-876 and TBI-223

The results for dose reproducibility are provided in Table 4. Average potency of three suspensions from three replicate aliquots of 2-mL from each suspension were found to be 98.9-99.7% for TBI-223 suspensions, and 102.6-103.2% for TBAJ-876 suspensions. The potency of an additional aliquot withdrawn after redispersing after 24 hours from each of the three suspensions was found to be 96.0-100.3% for TBAJ-876 suspensions and 99.3-100.3% for TBI-223 suspensions.

**Table 4.**
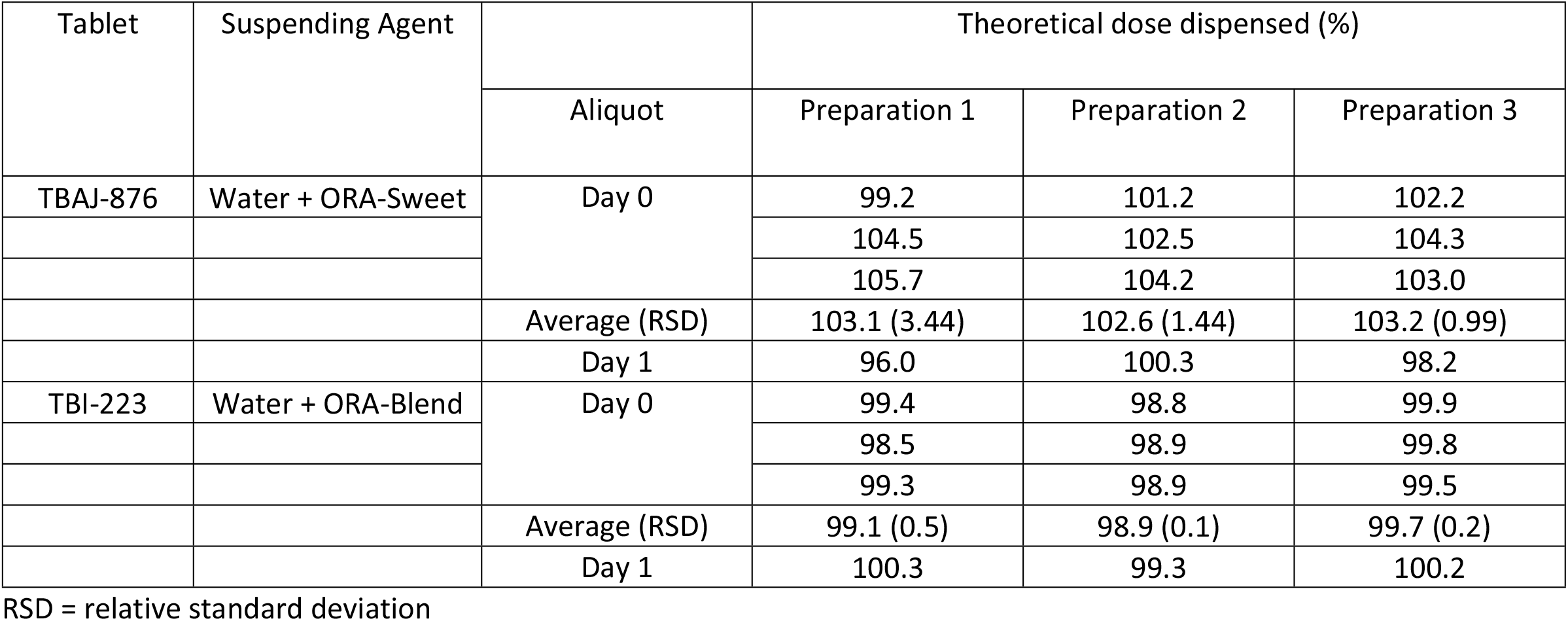
Dose Reproducibility of Aliquots Withdrawn from TBAJ-876 and TBI-223 Suspensions.

| Tablet | Suspending Agent |  | Theoretical dose dispensed (%) |  |  |
| --- | --- | --- | --- | --- | --- |
|  |  | Aliquot | Preparation 1 | Preparation 2 | Preparation 3 |
| TBAJ-876 | Water + ORA-Sweet | Day 0 | 99.2 | 101.2 | 102.2 |
|  |  |  | 104.5 | 102.5 | 104.3 |
|  |  |  | 105.7 | 104.2 | 103.0 |
|  |  | Average (RSD) | 103.1 (3.44) | 102.6 (1.44) | 103.2 (0.99) |
|  |  | Day 1 | 96.0 | 100.3 | 98.2 |
| TBI-223 | Water + ORA-Blend | Day 0 | 99.4 | 98.8 | 99.9 |
|  |  |  | 98.5 | 98.9 | 99.8 |
|  |  |  | 99.3 | 98.9 | 99.5 |
|  |  | Average (RSD) | 99.1 (0.5) | 98.9 (0.1) | 99.7 (0.2) |
|  |  | Day 1 | 100.3 | 99.3 | 100.2 |

RSD = relative standard deviation

#### TBAJ-876 tablets test method validation

The seven-point calibration curve in the range of 20 μg/mL to 60 μg/mL was linear with a high correlation coefficient (r) of 1.000. For both standard and sample tests, % RSD for inter- and intra-day precision were well below 2%, and mean recovery was well within the range of 98% to 102%.

#### TBI-223 tablets test method validation

The five-point calibration curve in the range of 25 μg/mL to 75 μg/mL was linear with a high correlation coefficient (r) of 1.000. For both standard and sample tests, % RSD for precision were well below 2%, and mean recovery was well within the range of 98% to 102%.

### Dispersions of commercial tablets prepared in XTEMP-R

The suspensions prepared with 5-minute soaking and 2-minute massaging times in the XTEMP-R device passed through the #18 sieve. No particles were retained indicating that all particles were below 1000 μm and dispersed particles were not forming aggregates or agglomerates.

The earliest dispersion times for all commercial tablets tested ranged from 1-3 minutes except for metformin which did not disintegrate within the time tested (10 minutes soaking and 5 minutes massaging). The dispersion times presented in Table 5 represent the dispersion time for the suspensions prepared by two independent analysts that passed the fineness of dispersion test.

**Table 5.** Dispersion Times of Commercial Tablets.

| Tablet | Dispersion Time (min) in X-TEMP-R |
| --- | --- |
| Valsartan | 1.5 |
| Acetaminophen | 3 |
| Glimepiride | 3 |
| Amlodipine | 1 |
| Metformin | >10* |

\* Did not disperse in time tested (10 minutes soaking and 5-minute massaging)

The drug content (labelled dose of the drug in total tablet weight) and disintegrant in each tablet formulation are provided in Table 2. Except for metformin, all other tablet formulations contained superdisintegrants. Valsartan, acetaminophen, and metformin had 31, 82, and 93% of drug load, respectively. Glimepiride and amlodipine contained 1.2 and 2.5% of drug load, respectively.

## Discussion

Tablet splitting, crushing, and grinding is a common practice for medication administration. The necessity for splitting or crushing tablets arises from the need to administer drug doses other than that are available in tablet formulations, as well as facilitate the swallowing of tablets [11]. There have been some challenges identified with both, tablet splitting and tablet pulverization. Large deviations of dose can occur due to unequal fragments of split tablets or weight differences due to loss of powdered or chipped material resulting from the splitting process [9]. Tablet crushing can result in significant drug loss [5]. Some crushing devices yield large particles that cannot be administered via a feeding tube [4]. Aerosolized powder during crushing and grinding can be an environmental [2] and health hazard.

This paper describes a procedure for converting tablets into a suspension: (1) that is simple and reproducible; (2) where tablet disintegration and preparation of suspension is performed within the same device; (3) in a contained environment; (4) that enables measurement and withdrawal of any required dose with accuracy; (5) with small particle size that permits administration through feeding tubes; and (6) where consumption is possible from the same receptacle.

XTEMP-R is a simple mechanical device designed to facilitate and expedite the disintegration and dispersion of tablets in a small volume (<10 mL) of water. This device is constructed with low hardness silicone, making it pliable yet durable. It can be gently squeezed by fingers to facilitate the dispersion of the tablet. This squeezing and massaging action facilitated the interaction of water with the superdisintegrants present in the tablet, thus promoting its dispersion. These ingredients are routinely included in the tablet formulations to aid in the break-up of the compacted mass.

The disintegration and suspension of tablets in the XTEMP-R device occurs in the same container in a closed environment. Therefore, there is minimal risk of any drug loss during the preparation. The XTEMP-R container does not have any cutting or grinding parts. It is also safer for caregivers to prepare the suspension with XTEMP-R since there is minimal contact with the drug either by touch or by inhalation of drug powder that may get aerosolized during pulverization.

Both TBAJ-876 and TBI-223 tablets are investigational drugs under IND applications with established specifications and content uniformity standards. As demonstrated by the results, TBAJ-876 and TBI-223 tablets could be dispersed homogenously in water and suspending vehicle within 10 minutes. Accurate aliquots could be withdrawn reproducibly from the device immediately after the preparation and 24 hours later. We withdrew aliquots from each preparation sequentially after preparation and on the following day after redispersing by shaking. This closely simulates what would occur in the pharmacy, at the patient’s home or at the clinical investigation site. The partial doses withdrawn for the suspension were uniform and stable as they could be redispersed reproducibly after 24 hours. It was not our intent to prepare suspensions for long term storage.

Fineness of dispersion was one of the quality criteria. The suspensions were tested by passing them through a #18 sieve (1000 μm aperture). Our objective was to ensure that no particle agglomerates were present in the suspension and that the largest particles in the suspension could be suspended in commonly available suspending vehicles. Bowles and colleagues [13] have demonstrated that particles up to 1000 μm can be successfully suspended with the aid of commonly used suspending vehicles.

Drug loss is one of the challenges when the tablets are pulverized before dispersing in a suspension. Drug powder tend to stick to the walls of the container such as a mortar and pestle. Thong and colleagues [5] have recommended two rinses of the tablet crushing devices to ensure minimum drug loss. Close to 100% of active drug from all three suspensions of TBAJ-876 and TBI-223 prepared by the XTEMP-R device was recovered. The devices were only rinsed once with 15 mL of water for each preparation. The total dose could be ingested directly from the device avoiding a transfer to a dispensing container. This methodology offers an alternative dose administration option for patients suffering from dysphagia.

We have also investigated the applicability of this device for some commercial tablets. Except for metformin, the tablets could be dispersed by soaking for 5 minutes and massaging for 2 minutes. For some products, the dispersion could be faster by just massaging for 1-3 minutes. The metformin tablets had a high drug load with just about 8% excipients and did not contain any superdisintegrants. Desai and colleagues [14] studied metformin formulation with minimal excipients and concluded that even for a highly soluble drug, a superdisintegrant was needed in the formulation to change the dissolution mechanism from erosion-diffusion to disintegration. Masum and colleagues [15] formulated metformin tablets with different disintegrants and found that the wetting time, dispersion time and the disintegration time of metformin tablets were governed by the type and quantity of the superdisintegrants. In the XTEMP-R device, the volume of the dispersing vehicle (5-7 mL water) and massaging are not sufficient to have effective erosion-diffusion for the tablet to dissolve or disperse the metformin tablet.

Therefore, the use of the XTEMP-R device may be limited for tablets that depend primarily on solubilization for dissolution and do not contain effective disintegrants. These observations corroborate our assumption that XTEMP-R promotes the interaction between water and tablet formulations. The ingredients of each tablet formulation can be found in the product prescribing information.

The lack of child-friendly formulations leaves 40% of the world population at increased risk for preventable adverse events, suboptimal dosing, noncompliance, and lack of access to new medicines [16]. In some instances, the caregiver is required to administer a quarter of a tablet to a neonate. XTEMP-R device may offer an alternative for accurate dosing in this patient population. Of course, appropriate development and analytical work will have to be performed.

Fortunately, several publications [17,18] describing the compatibility of drugs that lack pediatric formulations with commonly available suspending agents and with established storage conditions and shelf life are available.

### Limitations

XTEMP-R device worked well with the tablets containing superdisintegrants in the formulation. This device, however, may not work well with certain large tablets with a high drug-to-excipient ratio and/or tablets not containing effective disintegrants in the formulation. This will have to be tested on a case-by-case basis.

This device does not offer protection from light. Photosensitive drug preparations should be stored in the device with added protection from light.

The silicone material of the XTEMP-R device may be stained from a colored drug dispersion. Therefore, it is recommended that a dedicated device should be used for colored drugs such as clofazimine.

Dose preparation may be challenging for patients that are unable to massage the tablet in the device. XTEMP-E™, another version of XTEMP-R is equipped with an electronic mechanism for mixing. XTEMP-E will be presented in a subsequent publication.

The utility of this device and methodology for suspending any tablet formulation has to be supported by appropriate development and analytical work. As exemplified by the work conducted for TBAJ-876 and TBI-223 in this paper, similar rigor is recommended for any other drug that will utilize XTEMP-R for suspension.

## Conclusion

We have demonstrated that the new XTEMP-R device can be utilized for aliquoting partial doses of TBAJ-876 and TBI-223 immediate release tablets. Homogeneous suspensions could be prepared conveniently, safely, and expeditiously without any significant loss. Further, this device has the potential to offer an alternative for dosing patients who cannot ingest immediate release tablets. This could be beneficial to pediatric and elderly patients, intubated patients, and patients with dysphagia. Additional work is required to establish the utility of the device for other immediate release tablets.

## Data Availability

All relevant data are within the manuscript and its Supporting Information files.

## Acknowledgements

The authors are grateful for the funding provided by TB Alliance.

## Supporting Information

Link to video for suspension preparation methodology https://www.dropbox.com/sh/t0bj2o6ivjuh8l3/AAAZe3mRxc1HImGlbpoFDUUua?dl=0

**S1 Fig. Representative chromatograms of TBAJ-876 analysis** (a) Standard 1, (b) Diluent blank, (c) Day 0 aliquot for dose reproducibility, (d) Day 0 suspension matrix blank (diluent + ORA-Sweet), (e) Day 1 aliquot for dose reproducibility, (f) Day 1 suspension matrix blank (diluent + ORA-Sweet), (g) Standard 2, (h) Total dose recovery

**S2 Fig. Representative chromatograms of TBI-223 analysis** (a) Standard 1, (b) Diluent blank, (c) Day 0 aliquot for dose reproducibility, (d) Day 0 suspension matrix blank (diluent + ORA-Blend), (e) Day 1 aliquot for dose reproducibility, (f) Day 1 suspension matrix blank (diluent + ORA-Blend), (g) Standard 2, (h) Total dose recovery

## Author contributions

Rajneesh Taneja conceptualized and designed the device and experiments

Joseph Scarim conceptualized and designed the device and experiments

Poonam G Pande designed and coordinated the testing, corresponding author

Anthony Scarim conducted the analytical work

Milap C Nahata consulted on the utility of the device

Rita K Jew consulted on the utility of the device

Koteswara Inabathina coordinated the device manufacture

